# Explanation of Hand, Foot, and Mouth Disease Cases in Japan Using Google Trends Before and During the COVID-19: Infodemiology Study

**DOI:** 10.1101/2022.06.26.22276919

**Authors:** Qian Niu, Junyu Liu, Zixi Zhao, Miyu Onishi, Asuka Kawaguchi, Anuradhi Bandara, Keiko Harada, Tomoki Aoyama, Momoko Nagai-Tanima

## Abstract

**Background:** COVID-19 pandemic affected common disease infections, while the impact on hand, foot, and mouth disease (HFMD) is unclear. Google Trends data is beneficial in approximately real-time statistics and easily accessed, expecting to be used for infection explanation from information-seeking behavior perspectives. We aimed to explain HFMD cases before and during COVID-19 using Google Trends data.

**Methods:** HFMD cases were obtained from the National Institute of Infectious Disease, and Google search data from 2009 to 2021 was downloaded using Google Trends in Japan. Pearson correlation coefficients were calculated between HFMD cases and the search topic “HFMD” from 2009 to 2021. Japanese tweets containing “HFMD” were retrieved to select search terms for further analysis. Search terms were retained with counts larger than 1000 and belonging to ranges of infection sources, susceptible sites, susceptible populations, symptoms, treatment, preventive measures, and identified diseases. Cross-correlation analyses were conducted to detect lag changes between HFMD cases and HFMD search terms before and during COVID-19. Multiple linear regressions with backward elimination processing were used to identify the most significant terms for HFMD explanation.

**Results:** HFMD cases and Google search volume peaked around July in most years without 2020 and 2021. The search topic “HFMD” presented strong correlations with HFMD cases except in 2020 when COVID-19 outbroke. In addition, differences in lags for 73 (72.3%) search terms were negative, might indicating increasing public awareness of HFMD infections during the COVID-19 pandemic. Results of multiple linear regression demonstrated that significant search terms contained the same meanings but expanded informative search content during COVID-19.

**Conclusions:** Significant terms for HFMD cases explanation before and during COVID-19 were different. The awareness of HFMD infection in Japan may improve during the COVID-19 pandemic. Continuous monitoring is important to promote public health and prevent resurgence. Public interest reflected in information-seeking behavior can be helpful for public health surveillance.

## Background

Hand, foot, and mouth disease (HFMD) is an infectious disease that results in a blistering rash on the mouth, hands, and feet. Most infected individuals recover from HFMD within a few days. Various comorbidities, including myocarditis, neurogenic pulmonary edema, acute flaccid paralysis, and central nervous system complications, such as meningitis, cerebellar ataxia, and encephalitis, can also occur [1, 2]. HFMD has a worldwide distribution that outbreaks often occur during summer and early fall in the United States. Large outbreaks in Cambodia, China, Japan, Korea, Malaysia, Singapore, Thailand, and Vietnam have been reported in the past 2 decades [3, 4]. HFMD is seasonal in temperate Asia with a summer peak and subtropical Asia with spring and fall peaks, but not in tropical Asia, indicating a climatic role was identified for temperate Japan [5]. During the summer of 2011, Japan had the largest epidemic of HFMD on record, with 347,362 cases reported [6]. Coxsackievirus A6 (CV-A6) infection was responsible for most cases, with co-circulation of coxsackievirus A16 (CV-A16) and enterovirus A71 (EV-A71) [7]. EV-A71 has been sporadically detected from October 2014 onward. It became the predominant serotype in 2018, with approximately 70,000 reported cases, following an increased spread from the end of 2017 [8]. Since June 2019, a severe outbreak of HFMD has occurred in multiple regions of Japan, attracting public attention again [9]. As enteroviruses can spread rapidly by droplet and fomite transmission among children in daycare centers and kindergartens, understanding HFMD outbreaks is vital to public health, particularly during COVID [10].

Rapid recognition and reporting of HFMD infection are essential, and several studies have constructed models for explaining HFMD infection [11–15]. Rui et al. explored epidemiological characteristics and calculated the early warning signals of HFMD using a logistic differential equation (LDE) model in seven regions of China [11]. Yu et al. forecasted the number of HFMD cases with wavelet-based hybrid models in Zhengzhou, China [12]. Zhang et al. proposed a landscape dynamic network marker (L-DNM) to detect pre-outbreak signals of HFMD in Tokyo, Hokkaido, and Osaka, Japan [13]. Gao et al. used monthly HFMD infection cases and meteorological data to construct a weather-based early warning model with a generalized additive model across China [14]. Zhao et al. used a meta-learning framework and combines Baidu search queries for real-time estimation of HFMD cases [15]. The above studies used a range of data, including monthly or weekly HFMD infectious cases [11, 12, 14], dynamic information from city networks, horizontal high-dimensional data, records of clinic visits [13], meteorological data [14], and Baidu search queries data [15]. which are relatively difficult to access or delayed updates in Japan. Traditional surveillance and reporting systems lag an outbreak by one to two weeks because of the reporting and verification process. In Japan, the National Institute of Infectious Disease (NIID) has monitored the outbreak of various infectious diseases and issued weekly reports since 1999, but delayed for several weeks [16]. In addition, no studies have focused on changes in HFMD affected by the COVID-19 pandemic by Internet searching data compared with previous studies. As of August 2022, Google search data was considered reliable because its market share in Japan has been over 70% since 2009 [17, 18]. Google Trends data shows the information that the public is searching for more real-time and labor-saving, which may be valuable for infection surveillance.

The science of distribution and determinants of information in an electronic medium, specifically the Internet, or in a population, to inform public health and policy is defined as “infodemiology” [19]. Google Trends is frequently used in infodemiology research to gauge public interest [20]. Google Trends reflects public information-seeking behavior and allows users to analyze Google search data for specific search terms in any country or region over a selected period [21, 22]. Studies have shown that online query trends correlate with real-life epidemiologic phenomena such as the flu [23], sinusitis [24], lifestyle-related disease [25], asthma [26], and pruritus [27]. Researchers have also investigated public interest and information-seeking behavior in chronic obstructive pulmonary disease (COPD) [28], cancer [29, 30], bariatric surgery [31], kidney stone surgery [32], and suicide [33]. During the COVID-19 pandemic, similar studies using Google Trends search data were conducted to predict COVID-19 infectious cases [34, 35], explore public attitudes toward vaccination [36– 38], identify symptoms caused by pandemics [39–41], and assess affected medical services [42–44]. The above studies indicate that Google Trends could assist in gaining a better understanding and analysis of health information-seeking behavior. Information from Google Trends could be used to supplement the current infection reports with lag time.

This study aimed to explain HFMD infection using Google Trends data in Japan before and during the COVID-19 pandemic.

## Methods

### Data

We obtained actual HFMD cases from the weekly reports issued by NIID, which included new infectious cases and sentinel cases by prefecture and updated them from 1999 to the present [16]. Additionally, we set the geographic location to Japan and the category to health to limit irrelevant results and downloaded the relative search volume (RSV) of the “HFMD” search topic using Google Trends from January 1, 2009, to December 31, 2021. The normalized RSV data represented the search interest relative to the highest point for a given region and time. The scales of normalized RSV varied from 0 to 100, where 0 meant there were insufficient data for a term, while 100 was the peak popularity. We selected a search topic instead of the search term “HFMD” for comprehensive search information and limited the period from 2009 to 2021. In this study, we used the search topic “HFMD” and the search term “HFMD.” The weekly RSV of the search topic “HFMD” from 2009 to 2021 was gathered for further analysis.

To identify significant factors of HFMD infection, we created multiple linear regression models using HFMD-related search terms selected by Japanese tweets. We retrieved Japanese tweets through the publicly available Twitter Stream application programming interface (API) by querying the keywords “HFMD” to select “HFMD” related top words. Google applied improvements to the data collection system on January 1, 2016, and January 1, 2022, respectively. For consistency, 275,010 tweets restricting between 2016 and 2021 were downloaded in this study. Tokenization was used to select top words in Japanese tweets, which is a fundamental step in many natural language processing (NLP) methods, especially for languages like Japanese that are written without spaces between words. We tokenized all tweets and analyzed the unigram tokens. The website links, special characters, numbers, and “amp” (ampersands) were removed from the tweets before tokenization. The Python packages SpaCy and GiNZA were used to remove the Japanese stop words and implement tokenization. White space characters joined the tokenized words into text in the original order. The Python package scikit-learn was used to convert the white space–joined texts into unigram and bigram tokens and calculate the token counts. We provided counts of tokens in **Appendix 1**. Search terms with counts larger than 1000 and belonging to ranges of infection sources, susceptible sites, susceptible populations, symptoms, treatment, preventive measures, and identified diseases were selected for further analysis. Selected terms with corresponding interpretations and categories are provided in **Appendix 2** and **Appendix 3**. We downloaded the weekly RSV of selected search terms through Google Trends in two periods: before (2016-2019) and during the pandemic (2020-2021) based on the first case of COVID-19 in Japan was confirmed on 16 January 2020.

### Statistical Analysis

Initially, we calculated the Pearson correlation coefficient between the actual HFMD cases and RSV of the search topic “HFMD” each year from 2009 to 2021 instead of the whole period due to the periodic characteristic. Since our response variable HFMD cases and explanatory variables search term RSV are measured on a continuous scale, the parametric test should typically be selected instead of non-parametric analysis [45].

Second, we conducted cross-correlation analysis between actual HFMD cases and RSV of selected search terms. Cross-correlation is a measure of the similarity between two series as a function of the displacement of one relative to the other and was used to objectively estimate the time lag between cases of HFMD infection and related search terms [46]. We set the maximum lag to ±20 weeks due to the periodic characteristics of HFMD infection. We obtained 40 cross-correlation coefficients for each HFMD-related search term before and during the COVID-19 pandemic. Next, we selected the coefficients with the greatest absolute values and exhibited their true values. Finally, we compared the coefficients with the greatest absolute value in the periods before and during the COVID-19 pandemic. Regarding these coefficients, we assumed negative, zero, or positive values with the greatest absolute value representing the search terms that occurred earlier, coincided with, or later than the actual HFMD cases. Differences in lags during and before the COVID-19 pandemic was calculated to determine public awareness of HFMD.

Third, we conducted multiple linear regression to identify the most important Google search terms for explaining HFMD infection before and during the COVID-19 pandemic. We included HFMD-related search terms for multiple linear regression explanatory variables, with actual HFMD cases as response variables. Collinearity is the correlation between explanatory variables that expresses a linear relationship in a regression model. When the explanatory variables are correlated in the same regression model, they cannot explain the response variable dependently. We normalized the RSV of each selected term to avoid collinearity in regression models. Several common methods were used for explanatory variable selection to identify the most significant search terms and limit the number of explanatory variables, including forward selection, backward elimination, and stepwise regression [47]. Backward elimination was used in this study to find the best subset of search terms due to its easy implementation and automated availability. We used a p-value threshold of 0.05 to remove unnecessary search terms. In each round, we removed the search term with the highest p-value and reconstructed a multiple linear regression model until all p-values were under the threshold.

To assess the performance of the linear regression model, the coefficients of determination R^2^ or adjusted R^2^, which indicates how much variation in response is explained by the model, are often used [48]. We selected the adjusted R^2^ value for model evaluation to avoid overfitting the model.

## Results

### Basic description of HFMD cases and “HFMD” RSV

**Figure 1** presents the actual HFMD and RSV cases from 2009 to 2021. Visual inspection of the figure indicated that both the actual HFMD cases and RSV of Google Trends peaked around July in most years except for 2020 and 2021. The number of HFMD infections surged after 2011, peaking every two years before 2020. The RSV coincided with this trend. In 2020, no periodic peak of infection was observed, whereas, in 2021, the peak was delayed to November.

**Figure 1.**
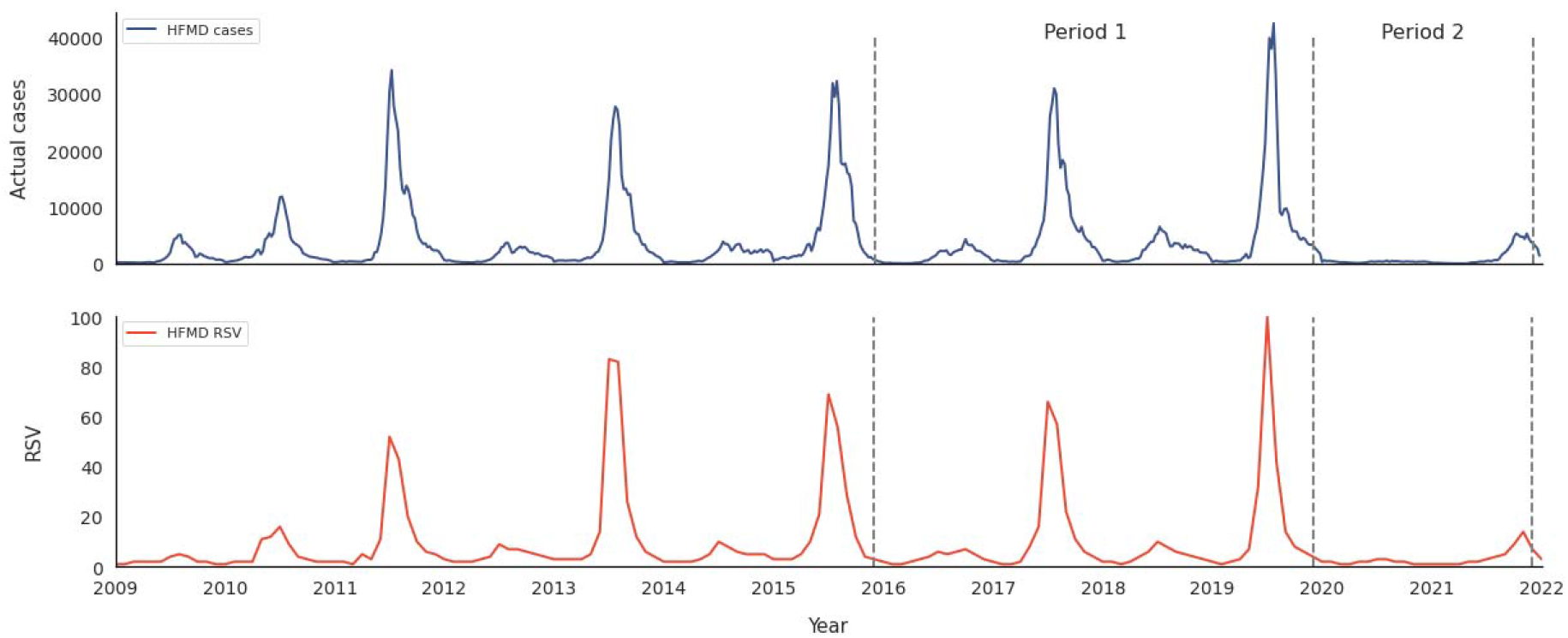
Monthly number and RSV of hand, foot, and mouth disease from 2004 to 2021.

As shown in **Figure 2**, we calculated the correlation between the actual HFMD cases and RSV each year from 2009 to 2021. These correlations’ mean (standard error) was 0.820 (0.052). Most coefficient values were greater than 0.7, except for 0.338 in 2020. Pearson correlation coefficients were provided in **Appendix 4**.

**Figure 2.**
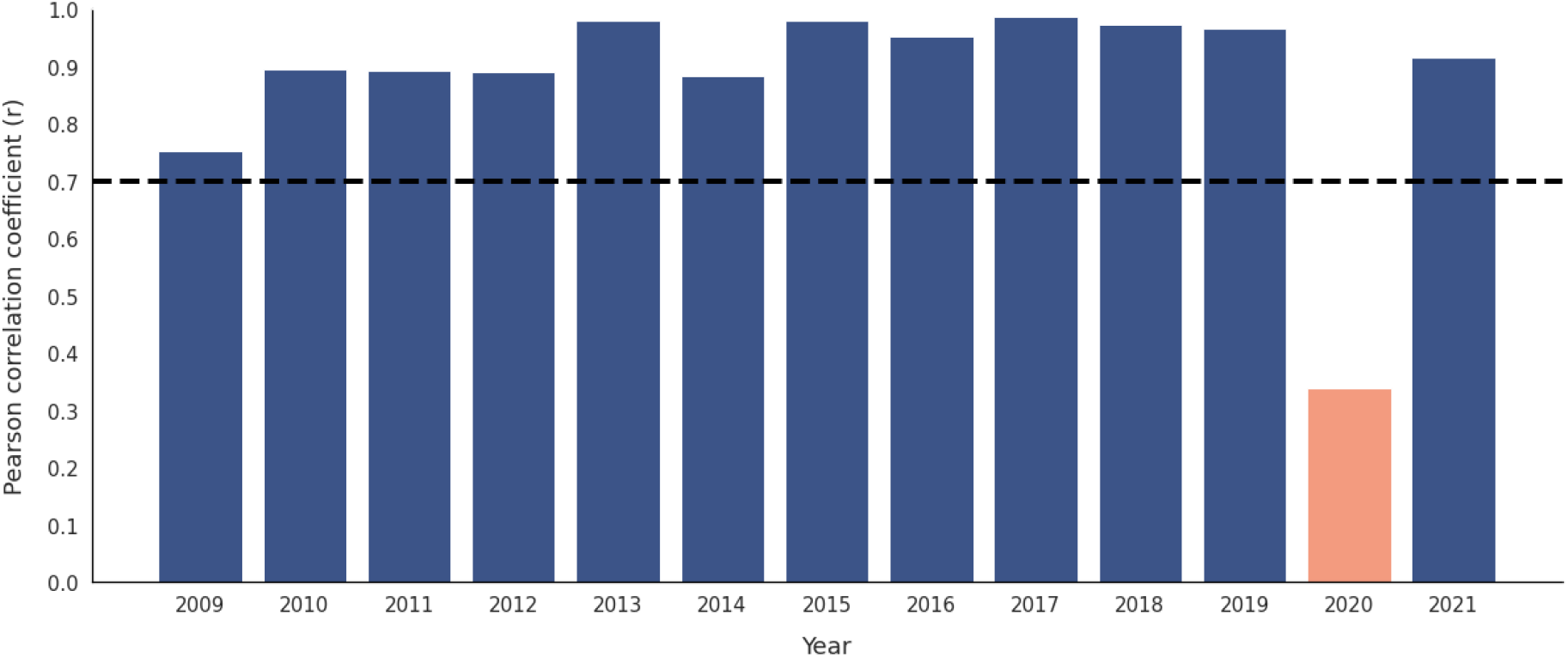
Correlation between the hand, foot, and mouth disease cases and the RSV from 2004 to 2021.

### Cross-correlation between HFMD cases and search term RSV before and during the pandemic

We performed cross-correlation analysis to determine the temporal relationship between HFMD cases and search term RSV. Cross-correlation results before and during the pandemic were presented in **Table 1**.

**Table 1.**
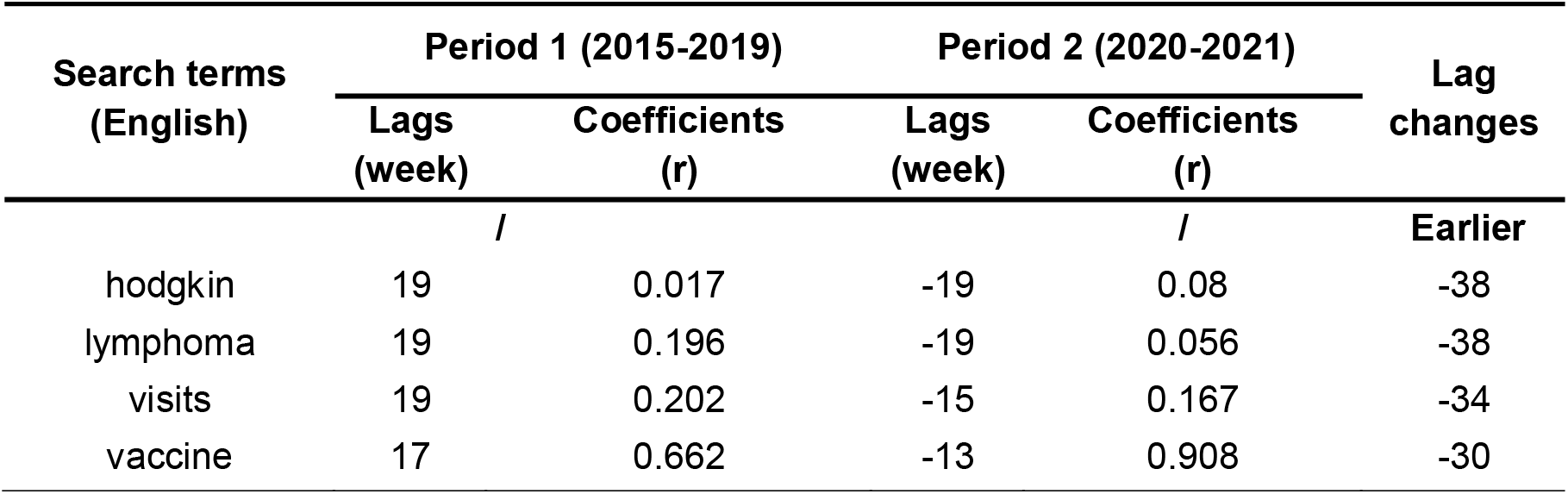
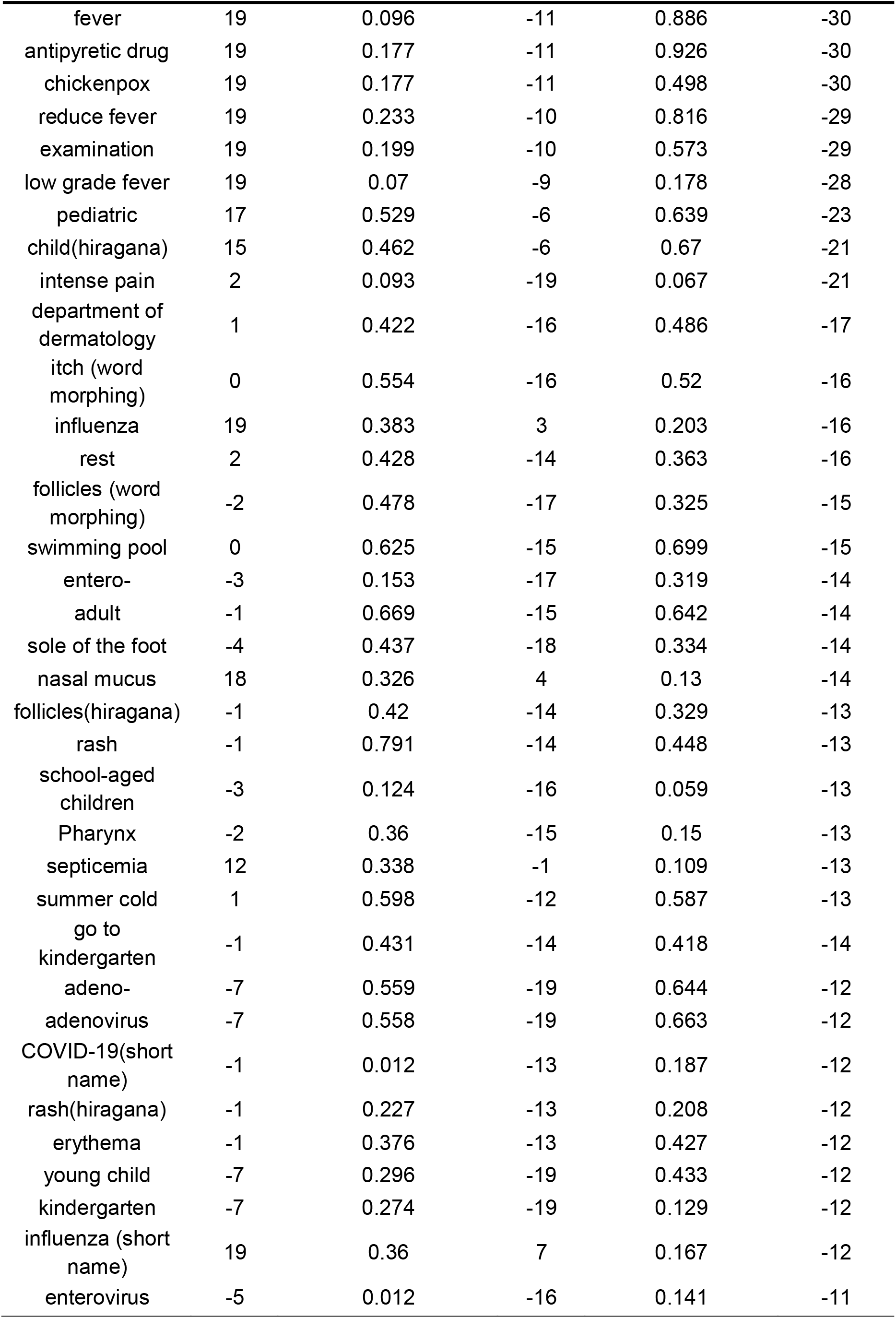
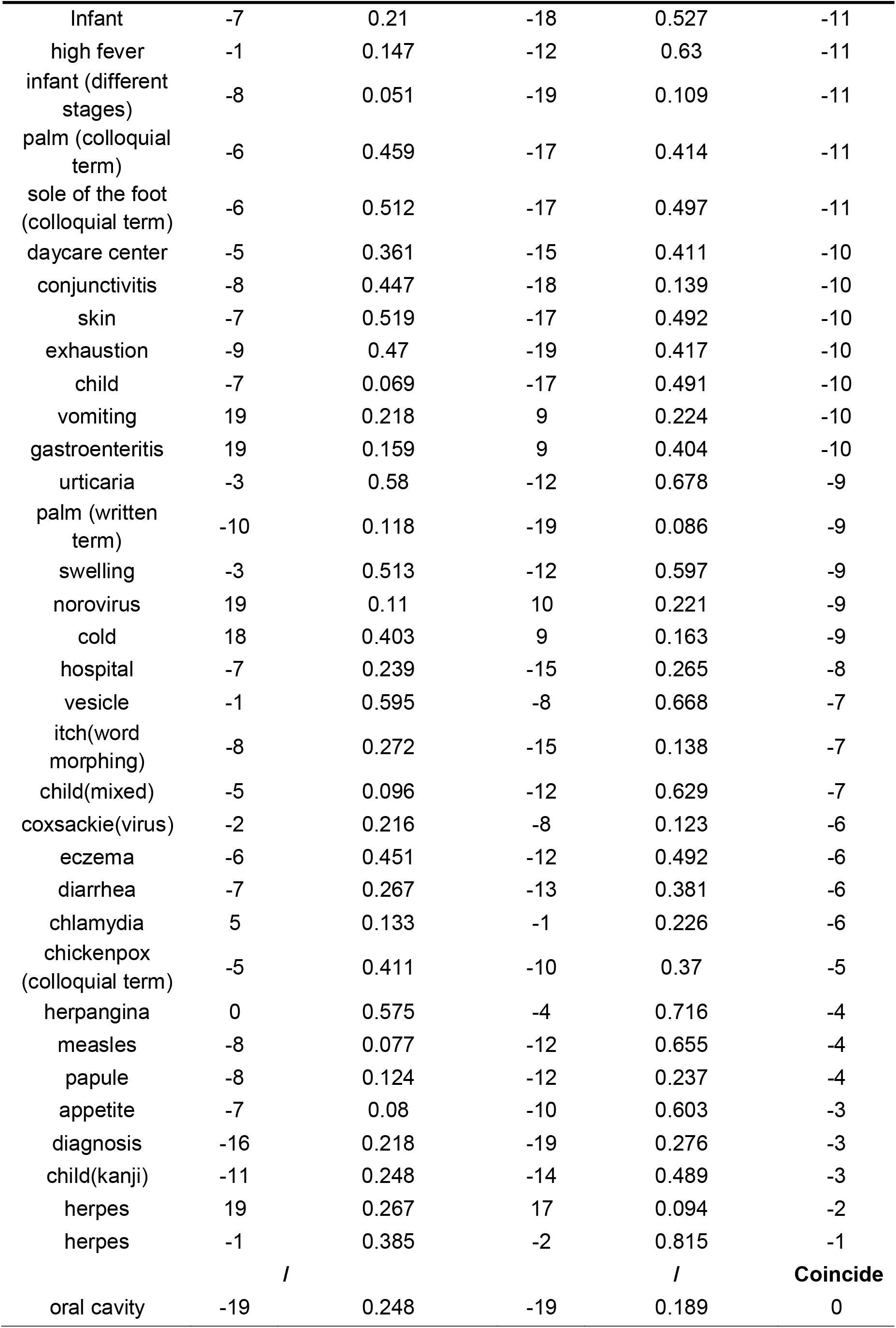
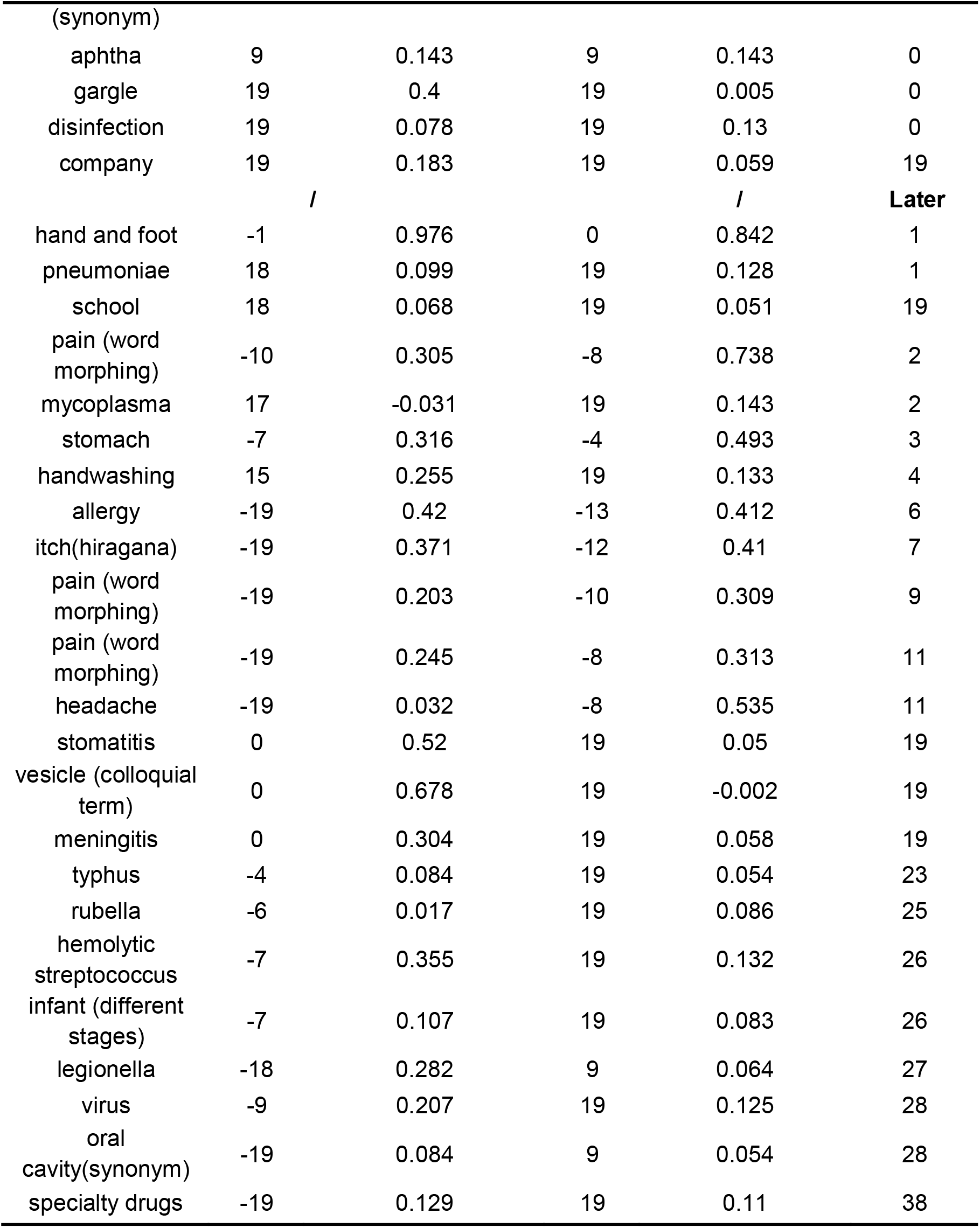
Cross-correlation analysis of HFMD cases and search terms RSV

Compared with period 1, the temporal correlation between HFMD cases and search term RSV changed in period 2. In period 1, 61 (60.4%), 6 (5.9%), and 34 (33.7%) of search term RSV presented earlier, coincide, and later than HFMD cases. In period 2, 73 (72.3%), 1 (1%), and 27 (26.7%) presented earlier, coincide, and later than HFMD cases. Differences in lags for 73 (72.3%) search terms were negative, might indicating increasing public awareness of HFMD infections during the COVID-19 pandemic. In contrast, lags for 5 search terms had no change, and 23 search terms exhibited delays.

### Essential search terms for explaining HFMD cases before and during the pandemic

We identified the most significant search terms for HFMD infection using multiple linear regression with backward elimination procedures. As shown in **Table 2**, 18 search terms were significant in period 1 and accounted for an adjusted R^2^=96.7% of the variation in HFMD infection. Conversely, as shown in **Table 3**, 57 search terms were detected as significant with adjusted R^2^=98.4% in period 2, which included more critical variables than in period 1. Model specification formulas were provided in **Appendix 5**.

**Table 2.** Multiple linear regression model of the hand, foot, and mouth disease cases and relative search volume of selected search terms from 2016 to 2019

| Search terms (English) | Model summary |  |  |
| --- | --- | --- | --- |
| | Standardized $\beta$ | Std.Error | P-value |
| virus | 0.0507 | 0.019 | 0.007 |
| herpangina | -0.1034 | 0.033 | 0.002 |
| diarrhea | -0.0998 | 0.021 | <0.001 |
| oral cavity (synonym) | 0.0290 | 0.015 | 0.049 |
| summer cold | 0.1361 | 0.037 | <.001 |
| adult | 0.1306 | 0.032 | <.001 |
| young child | -0.0372 | 0.017 | 0.032 |
| pediatric | 0.0641 | 0.019 | 0.001 |
| hand and foot | 0.8575 | 0.026 | <0.001 |
| chickenpox (colloquial term) | -0.0634 | 0.018 | 0.001 |
| eczema | 0.0506 | 0.018 | 0.006 |
| intense pain | 0.0361 | 0.014 | 0.010 |
| exhaustion | -0.0474 | 0.021 | 0.024 |
| itch (word morphing) | 0.0468 | 0.019 | 0.014 |
| swelling | -0.0481 | 0.022 | 0.033 |
| reduce fever | -0.1042 | 0.021 | <0.001 |
| sole of the foot | -0.0589 | 0.023 | 0.012 |
| nasal mucus | -0.0743 | 0.023 | 0.001 |

1. Adjusted $R^2 = 96.7\%$

**Table 3.** Multiple linear regression of the hand, foot, and mouth disease cases and relative search volume of selected search terms between 2020 and 2021

| Search terms (English) | Model summary |
| --- | --- |

| | Standardized<br>$\beta$ | Std.Error | P-value |
| --- | --- | --- | --- |
| adenovirus | -0.2816 | 0.04 | <0.001 |
| influenza (short name) | -0.1938 | 0.062 | 0.003 |
| entero- | 0.1566 | 0.021 | <0.001 |
| stomach | 0.0897 | 0.028 | 0.002 |
| coxsackie (virus) | 0.0446 | 0.02 | 0.034 |
| child (hiragana) | -0.4402 | 0.111 | <0.001 |
| COVID-19 (short name) | 0.6273 | 0.102 | <0.001 |
| follicles(katagana) | 0.1868 | 0.029 | <0.001 |
| herpangina | 0.8604 | 0.044 | <0.001 |
| herpes | -0.3042 | 0.042 | <0.001 |
| hodgkin | 0.0464 | 0.017 | 0.009 |
| mycoplasma | 0.1905 | 0.069 | 0.008 |
| legionella | 0.1888 | 0.025 | <0.001 |
| vaccine | -1.1388 | 0.136 | <0.001 |
| diarrhea | 0.2803 | 0.056 | <0.001 |
| papule | 0.0863 | 0.02 | <0.001 |
| infant (different stages) | -0.2984 | 0.034 | <0.001 |
| infant (different stages) | 0.0712 | 0.024 | 0.005 |
| daycare center | 0.3336 | 0.049 | <0.001 |
| visits | 0.1898 | 0.038 | <0.001 |
| oral cavity (synonym) | 0.1055 | 0.034 | 0.004 |
| pharynx | 0.1182 | 0.022 | <0.001 |
| vomiting | -0.0955 | 0.046 | 0.045 |
| summer cold | -0.9272 | 0.061 | <0.001 |
| child (mixed) | 0.1727 | 0.045 | <0.001 |
| child (kanji) | 0.2808 | 0.041 | <0.001 |
| young child | -0.2281 | 0.049 | <0.001 |
| pediatric | 0.6066 | 0.114 | <0.001 |
| child | 0.1418 | 0.045 | 0.003 |
| kindergarten | -0.2225 | 0.043 | <0.001 |
| palm (colloquial term) | 0.1388 | 0.029 | <0.001 |
| handwashing | -0.1137 | 0.051 | 0.031 |
| septicemia | 0.1228 | 0.017 | <0.001 |
| vesicle (colloquial term) | -0.2038 | 0.037 | <0.001 |
| vesicle | 0.1718 | 0.035 | <0.001 |
| chickenpox | 0.0902 | 0.021 | <0.001 |
| disinfection | -0.4493 | 0.082 | <0.001 |
| hemolytic streptococcus | 0.3638 | 0.086 | <0.001 |
| specialty drugs | -0.4039 | 0.047 | <0.001 |
| herpes | 0.1362 | 0.042 | 0.002 |
| exhaustion | -0.3952 | 0.041 | <0.001 |
| hospital | -0.214 | 0.05 | <0.001 |
| itch (word morphing) | 0.1993 | 0.032 | <0.001 |
| pain (word morphing) | -0.2462 | 0.047 | <0.001 |
| pain (word morphing) | -0.2206 | 0.029 | <0.001 |
| rash | -0.2028 | 0.055 | 0.001 |
| skin | 0.1754 | 0.04 | <0.001 |
| conjunctivitis | -0.1782 | 0.035 | <0.001 |
| pneumoniae | 0.3648 | 0.06 | <0.001 |
| swelling | 0.152 | 0.043 | 0.001 |
| urticaria | 0.747 | 0.147 | <0.001 |
| antipyretic drug | 0.3263 | 0.105 | 0.003 |
| infant | 0.2101 | 0.035 | <0.001 |
| sole of the foot (colloquial term) | -0.2455 | 0.052 | <0.001 |
| high fever | 0.1784 | 0.07 | 0.014 |
| measles | -0.5895 | 0.126 | <0.001 |
| nasal mucus | -0.2792 | 0.05 | <0.001 |

1. Adjusted R2 = 98.4%

Compared with period 1, significant search terms in period 2 contained the same meanings and expanded informative search content. “Herpangina,” “nasal mucus,” “exhaustion,” “diarrhea,” “summer cold,” “young child,” “pediatric,” and “swelling” occurred in the original form. “Pain,” “reduce fever,” “oral cavity,” “chickenpox,” “itch,” and “sole of the foot” occurred in morphing or synonym forms. “Virus” was replaced by more specific infection sources in period 2, such as “adenovirus,” “entero-,” “coxsackie,” “mycoplasma,” “legionella,” and “hemolytic streptococcus.” Correspondingly, “adult” was superseded by other terms for susceptible populations, such as “child” and “infant” (Omit synonyms). Search terms related to susceptible sites, preventive measures, and treatment also were identified in period 2, such as “daycare center,” “kindergarten,” “handwashing,” “disinfection,” “hospital,” and “specialty drugs.” Multiple linear regression results corroborated the cross-correlation results and indicated that public awareness of HFMD might increase during COVID-19.

## Discussion

This study presented trends and correlations in HFMD cases and RSV of “HFMD” from 2009 to 2021. Cross-correlation analyses were conducted between HFMD cases and search terms RSV before and during the pandemic. Additionally, multiple linear regressions were used to identify the significant search terms for explaining HFMD cases in two periods. Our results indicated that HFMD cases and RSV peaked around July in most years, except in 2020 and 2021, and surged after 2011 with peaks every two years before 2020. The search topic “HFMD” exhibited strong correlations with HFMD cases except in 2020, when COVID-19 outbroke. Furthermore, cross-correlation and multiple regression results revealed that the public might have improved awareness of HFMD infection during the pandemic. To our knowledge, this study is the first to explain HFMD cases using Google search data and examine changes in information-seeking behavior towards HFMD affected by the COVID-19 pandemic. Google search data could supplement public health surveillance and help authorities respond to infectious diseases rapidly.

From 2009 to 2021, the RSV of “HFMD” coincided with the HFMD cases except in 2020, which showed similar trends and peaks. In Japan, HFMD peaks generally occur around July [5]. During the COVID-19 pandemic, different from previous HFMD peaks disappeared in 2020 and lagged to November 2021. In 2020, Google Trends search data did not match the “HFMD” cases, with a relatively small peak in July. Despite a small peak in the RSV of search topic “HFMD”, the volume was much lower than in previous years. In contrast, the Japanese government implemented several measures to control COVID-19 that might potentially influence the spread of HFMD. Respiratory droplets and contact routes were mainly infection routes in HFMD and COVID-19 [49, 50]. Therefore, the susceptible population of HFMD also stay safe by taking standard precautions during COVID-19, such as physical distancing, wearing a mask, regularly washing hands, and coughing into a bent elbow or tissue [51].

The global pandemic might have enhanced public awareness of HFMD in addition to COVID-19 from the evidence provided by our results. 73 (72.3%) search terms cross-correlated earlier with HFMD cases during COVID-19, and significant search terms detected in period 2 contain more informative information. Previous studies demonstrated that the prevalence of respiratory infectious diseases reduced during the COVID-19 pandemic, such as influenza, varicella, herpes zoster, rubella, and measles [52–58]. This might have been due to adherence to non-pharmaceutical interventions and lower non-polio enterovirus activity during the COVID-19 pandemic compared with 2014-2019 [59]. Switzerland had an unprecedented complete absence of pediatric enteroviral meningitis in 2020 [60]. In Japan, community-acquired pneumonia [61] and influenza [62] admissions have been reduced during the COVID-19 pandemic. COVID-19 preventative actions and better personal hygiene are beneficial for preventing the spread of diseases. However, the prevalence of common diseases may rise as the public gradually complies less with infection control measures in the upcoming season [61]. Consistent with our results, a peak in HFMD infection and public interest re-occurred in November 2021. Continuous monitoring of HFMD is required, and public information-seeking behavior may be helpful in public health surveillance.

Google search data was applied in our study to explain HFMD cases affected by COVID-19 instead of Baidu search data was used for real-time estimation of HFMD cases in China [15] and other categories of data for HFMD prediction or explanation [11–14]. Differing from previous studies, we paid attention to the distinction between the information-seeking behavior of HFMD before and during COVID-19 and attempted to explain the HFMD cases using Google Trends data which has the most market share in Japan [17]. Many researchers have shown that the Google search data represents the public interest in a specific topic. However, Google search data should be used cautiously as a surveillance system because large events can easily interfere with it. Combing fine-grained data like mobility data could help develop surveillance systems that can effectively exclude biased or irrelevant information to respond rapidly [63].

## Conclusion

This study described trends and correlations in HFMD cases with RSV of “HFMD” and identified significant search terms to explain HFMD infections before and during the COVID-19 pandemic. We found the prevalence of HFMD was abnormal during COVID-19, and public might enhance awareness of HFMD infection affected by the pandemic. It is critical to continuously monitor resurgent common infections as the public gradually reduces compliance with infection control measures. Public information-seeking behavior using Google search data may be useful for public health surveillance.

## Limitations

This study had several limitations. First, our findings are limited to those who used Google to search for health-related information, which may not represent the entire community. The results may be biased toward younger people, who are more digitally connected than older individuals, although the Google search engine market share in Japan is nearly 80% [17]. Second, Google Trends improved geographical assignment and data collection systems in 2011, 2016, and 2022. Our results in the basic description of HFMD cases and “HFMD” RSV might be affected by them. Third, search data analysis is hypersensitive to large events, so complementary instead of replacing traditional research methods. Fourth, the specific HFMD-related terms we selected by Twitter might not represent all search terms in public use, especially hiragana, katakana, kanji, and alphabets used in Japan. Fifth, we used search data from 2016 to 2019 to represent the period before the COVID-19 pandemic due to restrictions of Google Trends, which may not represent the entire period. Finally, during the processing of backward elimination for regression model construction, significant terms might be eliminated due to jointly insignificant. Although we can find out potential significant terms from all eliminated terms, it is unpracticable to conduct F-test multiple times because multiple hypothesis testing leads to lower confidence levels. Hence, we remained current processing results.

## Data Availability

All data produced in the present study are available upon reasonable request to the authors

## Abbreviations

HFMD: hand, foot, and mouth disease
EV-A71: enterovirus A71
CV-A6: coxsackievirus A6
CV-A16: coxsackievirus A16
NIID: National Institute of Infectious Disease
LDE: logistic differential equation
L-DNM: landscape dynamic network marker
COPD: chronic obstructive pulmonary disease
RSV: relative search volume

## Declarations

### Ethics Approval

All methods were carried out in accordance with relevant guidelines and regulations (DECLARATION OF HELSINKI). Ethical approval and consent to participate were not necessary as the study was based on openly available aggregated data.

### Consent for Publication

Not applicable

### Availability of Data and Materials

We used publicly available data published by Google Trends and the National Institute of Infectious Disease, Japan. All data generated or analyzed during this study are included in this published article (**Appendix 6**).

### Authors’ Contributions

QN, JL, MNT, and TA contributed to the study conception and design. QN, ZZ, AB, KH, MO, and AK collected data. QN, JL, and ZZ participated in data analysis and interpretation and drafted the manuscript. All authors contributed to the manuscript revision and approved the final version of the manuscript.

## Acknowledgments

I appreciate support by 2022-2023 Google PhD Fellowship.

## References

1. Huang J, Liao Q, Ooi MH, Cowling BJ, Chang Z, Wu P, et al. Epidemiology of recurrent hand, foot and mouth disease, China, 2008-2015. Emerg Infect Dis. 2018;24.

2. Wang J, Hu T, Sun D, Ding S, Carr MJ, Xing W, et al. Epidemiological characteristics of hand, foot, and mouth disease in Shandong, China, 2009–2016. Sci Rep. 2017;7:1–9.

3. Puenpa J, Wanlapakorn N, Vongpunsawad S, Poovorawan Y. The History of Enterovirus A71 Outbreaks and Molecular Epidemiology in the Asia-Pacific Region. J Biomed Sci. 2019;26:75.

4. Biggs HM. Hand, foot, & mouth disease - chapter 4 - 2020 Yellow Book. https://www.nc.cdc.gov/travel/yellowbook/2020/travel-related-infectious-diseases/hand-foot-and-mouth-disease. Accessed 26 Sep 2022.

5. Koh WM, Bogich T, Siegel K, Jin J, Chong EY, Tan CY, et al. The Epidemiology of Hand, Foot and Mouth Disease in Asia: A Systematic Review and Analysis. Pediatr Infect Dis J. 2016;35:e285–300.

6. Ministry of Health, Labour and Welfare. National Institute of Infectious Diseases. Infectious Disease Weekly Report (JAPAN IDWR). 2009;11.

7. Fujimoto T, Iizuka S, Enomoto M, Abe K, Yamashita K, Hanaoka N, et al. Hand, foot, and mouth disease caused by coxsackievirus A6, Japan, 2011. Emerg Infect Dis. 2012;18:337–9.

8. Kabele P, Mojhová M, Smíšková D. Hand-foot-mouth disease in puerperium. Ceska Gynekol. 2022;87:47–9.

9. IDWR 2019 No. 29 <Infectious Diseases to Watch> Hand, Foot, and Mouth Disease. 2014. https://www.niid.go.jp/niid/ja/hfmd-m/hfmd-idwrc/9017-idwrc-1929.html. Accessed 23 Aug 2022.

10. Sun BJ, Chen HJ, Chen Y, An XD, Zhou BS. The Risk Factors of Acquiring Severe Hand, Foot, and Mouth Disease: A Meta-Analysis. Can J Infect Dis Med Microbiol. 2018;2018:2751457.

11. Rui J, Luo K, Chen Q, Zhang D, Zhao Q, Zhang Y, et al. Early warning of hand, foot, and mouth disease transmission: A modeling study in mainland, China. PLoS Negl Trop Dis. 2021;15:e0009233.

12. Yu G, Feng H, Feng S, Zhao J, Xu J. Forecasting hand-foot-and-mouth disease cases using wavelet-based SARIMA–NNAR hybrid model. PLoS One. 2021;16:e0246673.

13. Zhang X, Xie R, Liu Z, Pan Y, Liu R, Chen P. Identifying pre-outbreak signals of hand, foot and mouth disease based on landscape dynamic network marker. BMC Infect Dis. 2021;21 Suppl 1:6.

14. Gao Q, Liu Z, Xiang J, Tong M, Zhang Y, Wang S, et al. Forecast and early warning of hand, foot, and mouth disease based on meteorological factors: Evidence from a multicity study of 11 meteorological geographical divisions in mainland China. Environ Res. 2021;192:110301.

15. Zhao Y, Xu Q, Chen Y, Tsui KL. Using Baidu index to nowcast hand-foot-mouth disease in China: a meta learning approach. BMC Infect Dis. 2018;18:398.

16. IDWR Surveillance Data Table 2022 week 18. 2022. https://www.niid.go.jp/niid/en/survaillance-data-table-english/11133-idwr-sokuho-data-e-2218.html. Accessed 23 Aug 2022.

17. Search Engine Market Share Japan. StatCounter Global Stats. https://gs.statcounter.com/search-engine-market-share/all/japan. Accessed 27 Sep 2022.

18. Search Engine Marketing Share around the World from US, Europe and Asia. Chandler Nguyen. 2008. https://www.chandlernguyen.com/blog/2008/11/29/search-engine-marketing-share-around-the-world-from-us-europe-and-asia/. Accessed 27 Sep 2022.

19. Eysenbach G. Infodemiology and infoveillance: framework for an emerging set of public health informatics methods to analyze search, communication and publication behavior on the Internet. J Med Internet Res. 2009;11:e11.

20. Mavragani A, Ochoa G, Tsagarakis KP. Assessing the Methods, Tools, and Statistical Approaches in Google Trends Research: Systematic Review. J Med Internet Res. 2018;20:e270.

21. Nuti SV, Wayda B, Ranasinghe I, Wang S, Dreyer RP, Chen SI, et al. The use of google trends in health care research: a systematic review. PLoS One. 2014;9:e109583.

22. Mavragani A, Ochoa G. Google Trends in Infodemiology and Infoveillance: Methodology Framework. JMIR Public Health Surveill. 2019;5:e13439.

23. Pervaiz F, Pervaiz M, Abdur Rehman N, Saif U. FluBreaks: early epidemic detection from Google flu trends. J Med Internet Res. 2012;14:e125.

24. Sharma D, Sandelski MM, Ting J, Higgins TS. Correlations in Trends of Sinusitis-Related Online Google Search Queries in the United States. Am J Rhinol Allergy. 2020;34:482–6.

25. Memon SA, Razak S, Weber I. Lifestyle Disease Surveillance Using Population Search Behavior: Feasibility Study. J Med Internet Res. 2020;22:e13347.

26. Mavragani A, Sampri A, Sypsa K, Tsagarakis KP. Integrating Smart Health in the US Health Care System: Infodemiology Study of Asthma Monitoring in the Google Era. JMIR Public Health Surveill. 2018;4:e24.

27. Tizek L, Schielein M, Rüth M, Ständer S, Pereira MP, Eberlein B, et al. Influence of Climate on Google Internet Searches for Pruritus Across 16 German Cities: Retrospective Analysis. J Med Internet Res. 2019;21:e13739.

28. Boehm A, Pizzini A, Sonnweber T, Loeffler-Ragg J, Lamina C, Weiss G, et al. Assessing global COPD awareness with Google Trends. Eur Respir J. 2019;53.

29. Schootman M, Toor A, Cavazos-Rehg P, Jeffe DB, McQueen A, Eberth J, et al. The utility of Google Trends data to examine interest in cancer screening. BMJ Open. 2015;5:e006678.

30. Phillips CA, Barz Leahy A, Li Y, Schapira MM, Bailey LC, Merchant RM. Relationship Between State-Level Google Online Search Volume and Cancer Incidence in the United States: Retrospective Study. J Med Internet Res. 2018;20:e6.

31. Linkov F, Bovbjerg DH, Freese KE, Ramanathan R, Eid GM, Gourash W. Bariatric surgery interest around the world: what Google Trends can teach us. Surg Obes Relat Dis. 2014;10:533–8.

32. Dreher PC, Tong C, Ghiraldi E, Friedlander JI. Use of Google Trends to Track Online Behavior and Interest in Kidney Stone Surgery. Urology. 2018;121:74–8.

33. Taira K, Hosokawa R, Itatani T, Fujita S. Predicting the Number of Suicides in Japan Using Internet Search Queries: Vector Autoregression Time Series Model. JMIR Public Health Surveill. 2021;7:e34016.

34. Husnayain A, Shim E, Fuad A, Su EC-Y. Predicting New Daily COVID-19 Cases and Deaths Using Search Engine Query Data in South Korea From 2020 to 2021: Infodemiology Study. J Med Internet Res. 2021;23:e34178.

35. Higgins TS, Wu AW, Sharma D, Illing EA, Rubel K, Ting JY, et al. Correlations of Online Search Engine Trends With Coronavirus Disease (COVID-19) Incidence: Infodemiology Study. JMIR Public Health Surveill. 2020;6:e19702.

36. Pullan S, Dey M. Vaccine hesitancy and anti-vaccination in the time of COVID-19: A Google Trends analysis. Vaccine. 2021;39:1877–81.

37. Diaz P, Reddy P, Ramasahayam R, Kuchakulla M, Ramasamy R. COVID-19 vaccine hesitancy linked to increased internet search queries for side effects on fertility potential in the initial rollout phase following Emergency Use Authorization. Andrologia. 2021;53.

38. An L, Russell DM, Mihalcea R, Bacon E, Huffman S, Resnicow K. Online Search Behavior Related to COVID-19 Vaccines: Infodemiology Study. JMIR Infodemiology. 2021;1:e32127.

39. Han J, Kamat S, Agarwal A, O’Hagan R, Tukel C, Owji S, et al. Correlation Between Interest in COVID-19 Hair Loss and COVID-19 Surges: Analysis of Google Trends. JMIR Dermatol. 2022;5:e37271.

40. Kardes S, Erdem A, Gürdal H. Public interest in musculoskeletal symptoms and disorders during the COVID-19 pandemic : Infodemiology study. Z Rheumatol. 2022;81:247–52.

41. Knipe D, Gunnell D, Evans H, John A, Fancourt D. Is Google Trends a useful tool for tracking mental and social distress during a public health emergency? A time-series analysis. J Affect Disord. 2021;294:737–44.

42. Cohen SA, Ebrahimian S, Cohen LE, Tijerina JD. Online public interest in common malignancies and cancer screening during the COVID-19 pandemic in the United States. Transl Res. 2021;7:723–32.

43. Akpan IJ, Aguolu OG, Kobara YM, Razavi R, Akpan AA, Shanker M. Association Between What People Learned About COVID-19 Using Web Searches and Their Behavior Toward Public Health Guidelines: Empirical Infodemiology Study. J Med Internet Res. 2021;23:e28975.

44. Adelhoefer S, Berning P, Solomon SB, Maybody M, Whelton SP, Blaha MJ, et al. Decreased public pursuit of cancer-related information during the COVID-19 pandemic in the United States. Cancer Causes Control. 2021;32:577–85.

45. Benesty J, Chen J, Huang Y, Cohen I. Pearson Correlation Coefficient. In: Cohen I, Huang Y, Chen J, Benesty J, editors. Noise Reduction in Speech Processing. Berlin, Heidelberg: Springer Berlin Heidelberg; 2009. p. 1–4.

46. Bourke. Cross correlation. Cross Correlation”, Auto Correlation—2D Pattern.

47. Tranmer, Elliot. Multiple linear regression. Cathie Marsh Centre for Census ….

48. Akossou, Palm. Impact of data structure on the estimators R-square and adjusted R-square in linear regression. Int J Math Comput.

49. Huang C, Wang Y, Li X, Ren L, Zhao J, Hu Y, et al. Clinical features of patients infected with 2019 novel coronavirus in Wuhan, China. Lancet. 2020;395:497–506.

50. CDC. Causes & transmission. Centers for Disease Control and Prevention. 2022. https://www.cdc.gov/hand-foot-mouth/about/transmission.html. Accessed 28 Sep 2022.

51. Coronavirus disease (COVID-19): How is it transmitted? https://www.who.int/news-room/questions-and-answers/item/coronavirus-disease-covid-19-how-is-it-transmitted. Accessed 28 Sep 2022.

52. Sakamoto H, Ishikane M, Ueda P. Seasonal Influenza Activity During the SARS-CoV-2 Outbreak in Japan. JAMA. 2020;323:1969–71.

53. Wu D, Liu Q, Wu T, Wang D, Lu J. The impact of COVID-19 control measures on the morbidity of varicella, herpes zoster, rubella and measles in Guangzhou, China. Immun Inflamm Dis. 2020;8:844–6.

54. Wu D, Lu J, Liu Y, Zhang Z, Luo L. Positive effects of COVID-19 control measures on influenza prevention. Int J Infect Dis. 2020;95:345–6.

55. Kies KD, Thomas AS, Binnicker MJ, Bashynski KL, Patel R. Decrease in Enteroviral Meningitis: An Unexpected Benefit of Coronavirus Disease 2019 (COVID-19) Mitigation? Clin Infect Dis. 2021;73:e2807–9.

56. Veiga ABG da, Martins LG, Riediger I, Mazetto A, Debur M do C, Gregianini TS. More than just a common cold: Endemic coronaviruses OC43, HKU1, NL63, and 229E associated with severe acute respiratory infection and fatality cases among healthy adults. J Med Virol. 2021;93:1002–7.

57. Li Q, Wang J, Lv H, Lu H. Impact of China’s COVID-19 prevention and control efforts on outbreaks of influenza. Biosci Trends. 2021;15:192–5.

58. Wan WY, Thoon KC, Loo LH, Chan KS, Oon LLE, Ramasamy A, et al. Trends in Respiratory Virus Infections During the COVID-19 Pandemic in Singapore, 2020. JAMA Netw Open. 2021;4:e2115973.

59. Kuo S-C, Tsou H-H, Wu H-Y, Hsu Y-T, Lee F-J, Shih S-M, et al. Nonpolio Enterovirus Activity during the COVID-19 Pandemic, Taiwan, 2020. Emerg Infect Dis. 2021;27.

60. Stoffel L, Agyeman PKA, Keitel K, Barbani MT, Duppenthaler A, Kopp MV, et al. Striking Decrease of Enteroviral Meningitis in Children During the COVID-19 Pandemic. Open Forum Infect Dis. 2021;8:ofab115.

61. Yan Y, Tomooka K, Naito T, Tanigawa T. Decreased number of inpatients with community-acquired pneumonia during the COVID-19 pandemic: A large multicenter study in Japan. J Infect Chemother. 2022;28:709–13.

62. Hirose T, Katayama Y, Tanaka K, Kitamura T, Nakao S, Tachino J, et al. Reduction of influenza in Osaka, Japan during the COVID-19 outbreak: a population-based ORION registry study. IJID Regions. 2021;1:79–81.

63. Arik SÖ, Shor J, Sinha R, Yoon J, Ledsam JR, Le LT, et al. A prospective evaluation of AI-augmented epidemiology to forecast COVID-19 in the USA and Japan. NPJ Digit Med. 2021;4:146.

